# PySIRTEM: An Efficient Modular Simulation Platform for The Analysis of Pandemic Scenarios

**DOI:** 10.1101/2025.05.02.25326889

**Authors:** Preetom Biswas, Giulia Pedrielli, K. Selçuk Candan

**Affiliations:** School of Computing & Augmented Intelligence Arizona State University, 699 S Mill Ave, Tempe, AZ 85281, USA

## Abstract

Conventional population-based ODE models struggle against increased level of resolution since incorporating many states exponentially increases computational costs, and demands robust calibration for numerous hyperparameters. `PySIRTEM` is a spatiotemporal SEIR-based epidemic simulation platform that provides high resolution analysis of viral disease progression and mitigation. Based on the authors-developed Matlab© simulator `SIRTEM`, `PySIRTEM` ‘s modular design reflects key health processes, including infection, testing, immunity, and hospitalization, enabling flexible manipulation of transition rates. Unlike `SIRTEM`, `PySIRTEM` uses a Sequential Monte Carlo (SMC) particle filter to dynamically learn epidemiological parameters using historical COVID-19 data from several U.S. states. The improved accuracy (by orders of magnitude) make `PySIRTEM` ideal for informed decision-making by detecting outbreaks and fluctuations. We further demonstrate `PySIRTEM` ‘s usability performing a factorial analysis to assess the impact of different hyperparameter configurations on the predicted epidemic dynamics. Finally, we analyze containment scenarios with varying trends, showcasing `PySIRTEM` ‘s adaptability and effectiveness.

## 1 INTRODUCTION

The devastating outbreak of COVID-19 disease, caused by the airborne SARS-CoV-2 virus, has had a long-lasting impact around the world, with 7 million deaths to date (Hossain et al. 2021). Throughout history, the human race has faced the doom of epidemics, pandemics, and outbreaks such as plague, flu, and Ebola, which pose a great threat to human life and economy. Various mitigation strategies have been employed by policymakers to curb the onslaught of infection such as lockdown, isolation, optimal vaccine allocation, preemptive quarantine, and random testing (Anderson et al. 2020; Walensky and Del Rio 2020; Roy et al. 2021). Despite recent significant breakthroughs in medical science, there are substantial challenges in impeding the destruction of pandemics. The incredible advancement of transportation, especially the ease of international air travel, allows for rapid dissemination of viruses with a high reproduction number such as COVID-19. The lack of reliable testing data for early detection of a novel virus outbreak poses an added obstacle. Acquiring reliable population-level epidemiological data is another challenging task due to possibilities of underreporting, misreporting, and testing limitations (Lau et al. 2021). In addition, the adaptive and mutative abilities of certain viruses lead to the inception of newer, often more virulent strains, stressing the need for a flexible and meticulous epidemiological model.

Duan et al. (2015) classify epidemiological models into three categories: complex network models, agent-based models, and analytical models. Network-based models focus on person-to-person interactions, representing disease spread as a space-time graph (Chang et al. 2021). Although limited by computational complexity for large-scale studies, they can be coupled with analytical models to infer inter-zonal relation and optimize containment policies. Here, each node acts as an individual zone with internal mobility and edges denote contagion influence between zones (Roy et al. 2021a). Agent-based models, which simulate individual behaviors and interactions, allow for high-resolution analysis of disease transmission. Various studies have employed agent-based modeling in the scope of COVID-19, Dengue, HIV, both independently and coupled with analytical models (Hoertel et al. 2020; Rhee 2006; Miksch et al. 2015; Frias-Martinez et al. 2011). Nevertheless, their use is also limited to smaller case studies due to high computational demands. Traditional analytical models such as differential equation-based SIR, SIS, SEIR, and SEIRS models are most commonly used to study population-level pandemic progression (Jit and Brisson 2011; Li and Muldowney 1995). To support policy optimization and decision making, these models can incorporate additional significant real-life events such as quarantine and hospitalization (He et al. 2020). For example, Chu et al. (2020) incorporated mask wearing, eye protection, and social distancing to study infection reduction while Eikenberry et al. (2020) embedded the effect of masks by adjusting infection rates. Moreover, testing campaigns play a crucial role in identifying outbreaks, tracking epidemic trends, and steering interventions, particularly in diseases like COVID-19 with significant asymptomatic spread. However, testing can be costly and unreliable. So epidemic models must address challenges such as cost constraints, daily capacity, test accuracy, sensitivity and specificity. As mass testing can be costly, choosing optimal testing strategies is significant in identifying target populations that require quarantine (Omori et al. 2020). Wells et al. (2021) also explored reducing the quarantine period with exit testing as an effective alternative to the 14-day full quarantine. Furthermore, embedding spatiotemporal mobility is another key factor in accurately predicting and simulating viral progression across zones and individuals (Niehus et al. 2020; Roy et al. 2021b).

While effective, any singular model fails to incorporate important health processes such as immunity, quarantine, hospitalization, and multiple testing modalities while accounting for population movement and viral evolution. In our earlier work (Azad et al. 2022), we introduced a spatially informed Susceptible-Exposed-Infected-Recovered-Susceptible (SEIRS) model, `SIRTEM`, which couples the spatio-temporal dynamics of the COVID-19 epidemic with above-mentioned compartments to solve the optimal cost-effective multi-modal COVID-19 testing strategy problem while satisfying practical constraints (Azad et al. 2022). In this paper, we present `PySIRTEM`, which implements and extends the `SIRTEM` model to accommodate dynamic learning of model parameters and provides flexible manipulation to simulate real-world viral interdiction scenarios accurately.

## Contributions

`PySIRTEM` builds upon the `SIRTEM` model with a novel Sequential Monte Carlo (SMC) particle filter approach that improves the accuracy in the implementation of transition hyperparameters. The black box nature of the particle filter allows for sensitive identification of rapid fluctuations, making `PySIRTEM` a powerful tool to capture intrinsic pandemic behavior. Additionally, we provide detailed analysis of the interaction between various model parameters and their impact on pandemic progression through Design of Experiments. Finally, we simulate multiple pandemic scenarios and analyze `PySIRTEM`’s effectiveness in informed decision and policy making.

## 2 APPROACH

### 2.1 SEIRS Epidemic Model

For `PySIRTEM`, we extend the Susceptible-Exposed-Infected-Recovered-Susceptible (SEIRS) model. Susceptible (S) population consists of individuals who are not exposed to infection. Once exposed, they may transition to the Exposed (E) class that denotes an asymptomatic or untested population. If tested positive, exposed individuals transfer to Infected (I) class from where they transition either to Recovered (R) or dead. SEIRS models further allow incorporation of waning immunity in disease modeling, where a recovered individual can again become susceptible over time. A set of differential equations regulate the transition between the health classes using parameters such as infection rate (*β*), recovery rate (*γ*), and loss of immunity rate (*δ*). However, the traditional SEIRS model fails to incorporate important dynamics and processes such as testing, hospitalization, quarantine, and natural and induced immunization.

**Figure 1.**
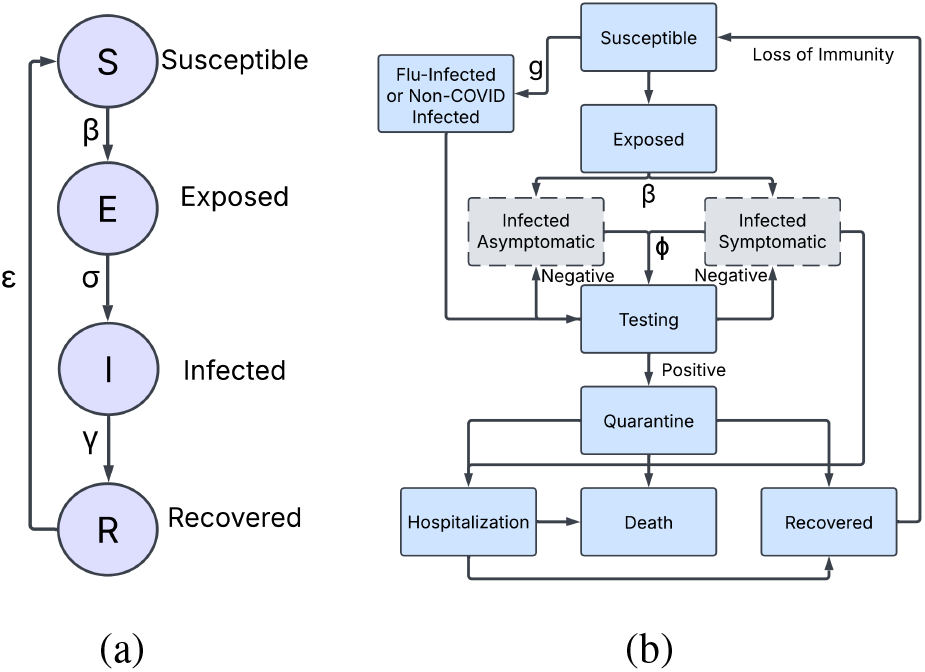
A brief overview of (a) SEIRS model and (b) `PySIRTEM` model

### 2.2 **PySIRTEM**

`PySIRTEM` implements the extended SEIRS model, `SIRTEM` (*Spatially Informed Rapid Testing for Epidemic Modelling*) (Azad et al. 2022), to simulate pandemic evolution. `PySIRTEM` builds upon the SEIRS model by incorporating additional states to account for testing, hospitalization, quarantine, and immunity loss. It also makes provisions for optimized symptomatic and asymptomatic testing strategies as well as the accuracy (false positive and false negative rate) of these test results. Furthermore, `PySIRTEM` is spatially informed i.e. accounts for the distribution of population in different zones and associated mixing rates.

`PySIRTEM` includes 46 medical states by dividing individuals into five population groups or compartments: *Susceptible* individuals who are non-infected and display no COVID symptoms; *Symptomatic Infected* individuals showing COVID symptoms; *Asymptomatic Infected* individuals who do not exhibit symptoms; *Symptomatic but not COVID-infected* individuals with flu-like symptoms; and *Falsely Presumed Susceptible* individuals who have natural immunity but are falsely tested negative for COVID antibodies. `PySIRTEM` also employs delay differential equations (DDEs) instead of traditional Ordinary Differential Equations to accommodate for delays in disease progression and testing results. Analogous to conventional SEIRS models, transition between the compartments are governed by relevant transition rates and parameters such as infection rate, testing rate, hospitalization rate and so on. Comprehensive overview of the subsequent processes and parameters are detailed in the SIRTEM paper (Azad et al. 2022).

### 2.3 Calibration of Transition Rates using Particle Filters

For a given geographic location, we calibrate `PySIRTEM` by estimating three critical transition parameters: infection rate (*β*), testing rate (*φ*), and general non-COVID-19 sickness rate (*g*) using published reported daily positive and daily negative cases. We employ particle filters, also known as Sequential Monte Carlo (SMC) methods, that are highly effective in approximating solutions of non-linear state space systems. Unlike traditional optimization approaches or Kalman filters, particle filters are non-parametric which are ideal for non-linear, non-Gaussian epidemic processes.

#### 2.3.1 Basics of a Particle Filter

Particle filters leverage a set of weighted particles to derive the posterior distribution of latent states in a state-space model. A state-space model consists of a latent Markov process (*X*_*t*_) and a series of noisy observations (*Y*_*t*_) at discrete time. The state transition model defines the evolution of the latent state and follows the following probability,

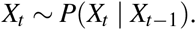

Observations are conditionally independent given the latent states and each observation is only dependant on the current latent state.

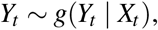

where *g*(·) denotes the observation likelihood function. Given a set of observations *Y*_1:*t*_ = {*Y*_1_,*Y*_2_, …, *Y*_*t*_}, the particle filter estimates the posterior distribution *π*(*X*_*t*_|*Y*_1:*t*_) by using a set of *N* weighted particles 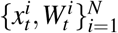 evolving over time, where *x*_*t*_ is a sampled particle from the distribution of *X*_*t*_. The weights are normalized such that 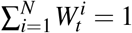. The particle filter recursively computes importance sampling approximations of *π*_*t*_ as follows,

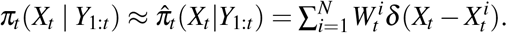

Here 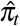 is the empirical approximation *π*_*t*_ using weighted particles and *δ* represents the Dirac delta function. The value of *δ* is 0 everywhere except at 0, and integrates to 1. The parameters *δ* is used to approximate the posterior distribution as the sum of weighted point masses. The algorithm consists of three major steps:

- **Resample:** Draw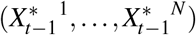 (with replacement) from 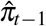 according to weights 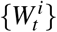 to discard lower weighted particle before propagation. Here 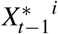 is a resampled particle.
- **Propagate:** Draw 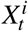 from 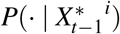, independently for different indices *i*.
- **Reweight:** Set 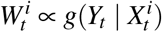.

Note that at time *t* = 0, particles are drawn from *π*_0_ and 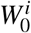 is set to 1*/N*. Further detailed discussions on particle filters can be found in (Künsch 2013; Doucet et al. 2009) and related literature.

#### 2.3.2 PySIRTEM State Space Model Setup And Particle Filter Iteration

In the context of epidemic modeling and forecasting, our goal is to estimate the following latent state vector:

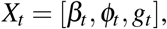

where, *β*_*t*_ is the infection rate for susceptible population, *φ*_*t*_ is the diagnostic testing rate for symptomatic individuals, and *g*_*t*_ is the ratio of susceptible population who have fever for non-COVID infections.

Given a latent space vector *X*_*t*_, `PySIRTEM` model produces two predicted time series Ŷ = [*ŷ*^+^, *ŷ*^−^] (predicted daily positives *ŷ*^+^ and negatives *ŷ*^−^), which are compared against the actual reported case numbers *Y* = [*ŷ*^+^, *ŷ*^−^] (reported daily positives *y*^+^ and negatives *y*^−^).

We assume *X*_*t*_ evolves in a non-linear, autoregressive manner and define the state transition and observation models as

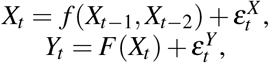

where *F* represents the `PySIRTEM` simulation function that takes in the transition rates and returns 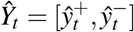. Here, both 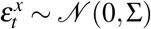 and 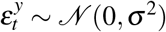 denote small random noise.

`PySIRTEM` utilizes a dynamic piecewise autoregressive order-2 (AR2) process to model the state transitions. We generate *N* = 200 initial particles 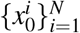 from a prior uniform distribution *U* (0, 1). In particle filter, approximation accuracy increases with the number of particles. Since large number of particles adds significant computational cost for `PySIRTEM`, we use an importance density (or proposal distribution) that incorporates the previous two predicted states (Equation 1, 2, 3), limiting search in relevant state space and providing better approximation with fewer particles (Arulampalam et al. 2002). For each particle 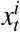, the state transition function is called and calculated using 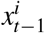 and 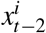 Only for 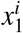, an AR(1) process is used.

The transition rates *x*_*t*_ = [*β*_*t*_, *φ*_*t*_, *g*_*t*_] are only updated weekly and kept constant otherwise. For a given week *k*, the rates are calculated as follows:

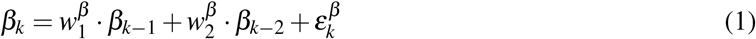

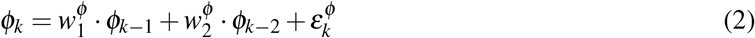

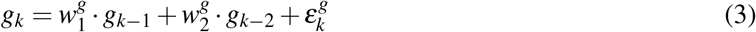

Above, the weights 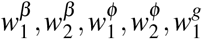, and 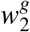 are randomly sampled between [0.1, 0.8] each week so that the model does not overfit by allowing unrealistic fluctuations while providing enough flexibility to reflect real-world changes quickly. A small Gaussian perturbation *ε*_*k*_ ~ *𝒩* (0, *σ* ^2^) is added for stochasticity. The new rates are bounded between [0, 1] to ensure realistic values.

The `PySIRTEM` model simulation is run using each of the state particles as input:

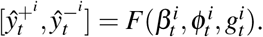

We compare the outputs with reported case data obtained from public sources (CTW nd) and update the weights by computing the following likelihood

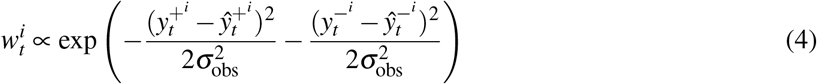

Here 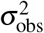 represents the noise in observational data. The weights are normalized and *N* particles are resampled at the next time point proportionate to their updated weights, but are only updated weekly. The weekly update is to ensure the model does not overfit and also to account for the delay periods in epidemic processes. The process is repeated throughout the horizon and the Maximum a Posteriori (MAP) estimation for *β*_*t*_, *φ*_*t*_, *g*_*t*_ are recorded at each timepoint.

### 2.4 Factorial Analysis

Factorial analysis (Montgomery 2017) is a powerful approach to identify latent variables and gauge relationships between a set of observed variables. It can either be utilized as a means for identifying underlying structure or a tool for testing a hypothesized structure. In `PySIRTEM`, we apply factorial analysis to study the effects of various trends in transition parameters. Specifically, we analyze how different infection rate (*β*), testing rate (*φ*), and general sickness rate (*g*) impact daily case numbers. We explore both the main effects, i.e., how each factor individually influences outcome, and interaction effects, i.e., how the rates jointly impact the case trends.

For `PySIRTEM`, we conduct a full factorial experiment by varying the three rates in three distinct temporal shapes:

1. increasing: the rate gradually increases over time.
2. decreasing: the rate gradually decreases over time.
3. single-peak: the rate follows a bell-curve trajectory, peaking at an intermediate timepoint.

Additionally, each shape has different bounds denoting the highest and lowest value. Specifically, we denote the bounds in three classes: high, medium, and low, which result in a total of nine unique setups for each shape. The specific bounds for the rates are listed in Table 1.

**Table 1.** Range of rates for different shapes and orders for factorial analysis.

| Rate | Shape | Order | Range |
| --- | --- | --- | --- |
| Infection Rate ($\beta$) | Increasing, Decreasing, Single-peak | High | [0.05, 0.99] |
|  |  | Medium | [0.05, 0.7] |
|  |  | Low | [0.05, 0.5] |
| Testing Rate ($\phi$) | Increasing, Decreasing, Single-peak | High | [0.05, 0.4] |
|  |  | Medium | [0.05, 0.3] |
|  |  | Low | [0.05, 0.2] |
| General Sickness Rate ($g$) | Increasing, Decreasing, Single-peak | High | [0.001, 0.05] |
|  |  | Medium | [0.001, 0.03] |
|  |  | Low | [0.001, 0.02] |

#### Main Effects Analysis

Main effect in factorial design refers to the influence of a singular independent variable on the dependent variable, ignoring the effects of other independent variables. The main effect gives insight into whether changing the level of one factor affects the outcome variable or not. The main effect of a factor *A* is denoted by,

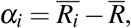

##### Algorithm 1

Particle Filter Iteration Overview

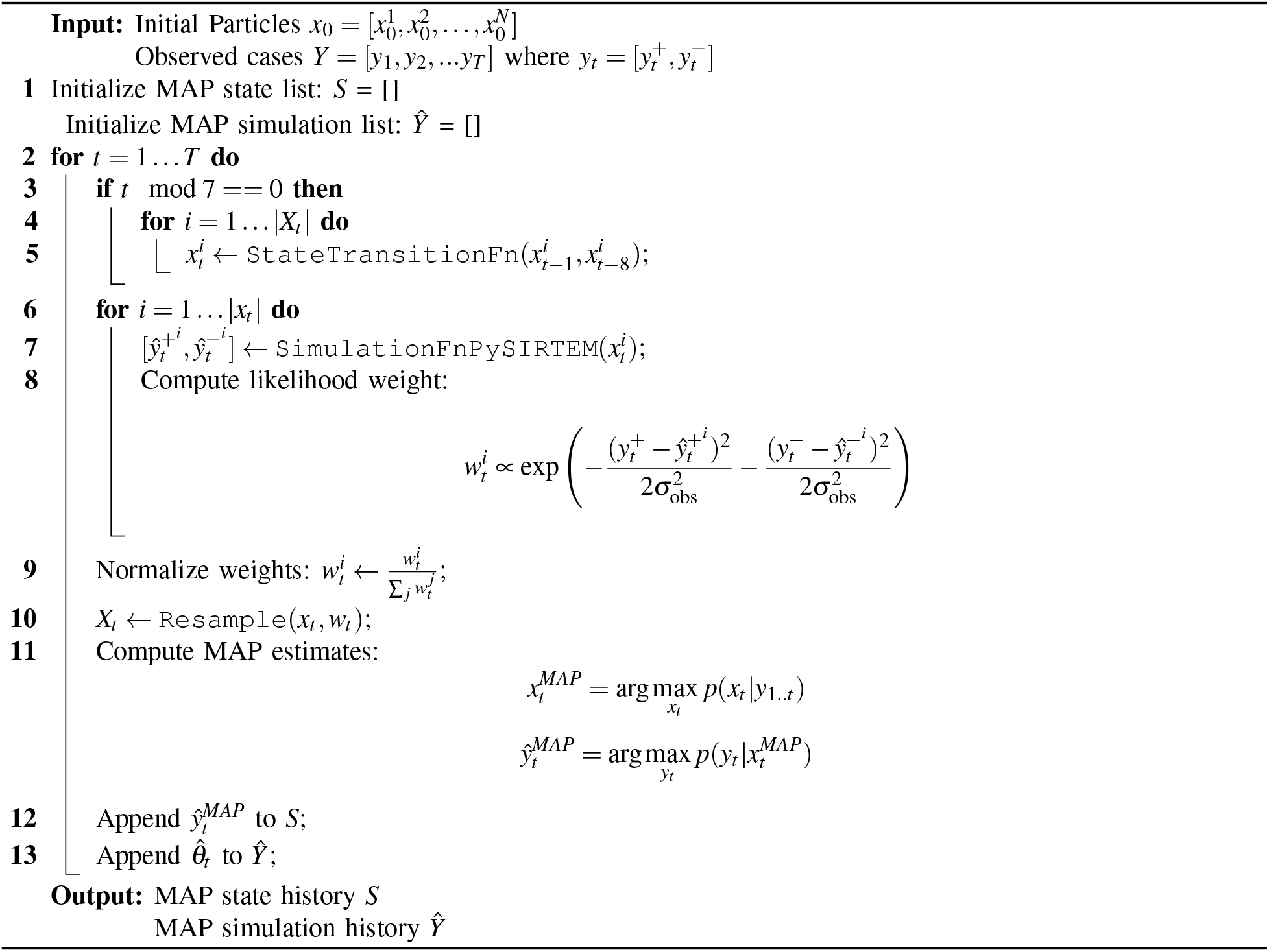

where 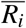 is the mean response for level *i* of factor *A* and 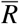 is the grand mean i.e the average response of all levels. In our experiment, we denote the levels of each factor by mapping the trends to 1 and −1 if they are used in the simulation or not respectively. For example, if a simulation is run with high increasing *β*, high increasing *φ*, and low single-peak *g*, we map these 3 factors to level 1 and all the other factors to level −1.

#### Interaction Plots Analysis

Interaction plots provide a visual report on how the relationship between one factor and the outcome depends on the levels of another factor and reveal additional insight of combined interactions possibly overlooked by main effects analysis. The interaction between level *i* of factor *A* and level *j* of factor *B* is denoted by

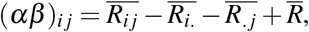

where 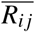 is the mean response for the combination of level *i* of *A* and level *j* of *B*, 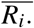 is the mean response for level *i* of *A* (ignoring *B*), 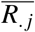 is the mean response for level *j* of *B* (ignoring *A*), and 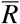 is the grand mean. A large positive or negative (*αβ*)_*i j*_ value indicates strong interaction while (*αβ*)_*i j*_ ≈0 means no interaction.

Parallel lines in the plot denote no interaction while non-parallel lines suggest one factor’s effect depends on the level of another factor. Crossing lines indicate strong interaction where the direction of the effect reverses based on the level of the other factor. Strong interaction between two factors imply that the effect of the factors cannot be evaluated in isolation and main effect analysis should be interpreted with caution.

## 3 EXPERIMENTAL RESULTS

### 3.1 PySIRTEM Validation

We confirmed the validity and accuracy of the `PySIRTEM` parameters through particle filter approach by comparing the the simulation results from the model against published confirmed case data from three

U.S. states - Arizona, Florida, and Minnesota. `PySIRTEM` is implemented in `Python 3.12.9` using libraries such as `NumPy`, `SciPy`, `ddeint`, and `pfilter` and run on Sol Supercomputer at Arizona State University, an environment with 12 CPUs and 8 GiB memory. The particle filter algorithm is initialized with *N* = 200 particles, with the infection rate (*β*), test rate (*φ*), and general sickness rate (*g*) for each state calibrated weekly. Weights were updated based on Equation 4 and importance sampling was employed to discard lower weighted particles. The full list of `SIRTEM` simulation parameters and their sources are detailed in the Azad et al. (2022) paper. `PySIRTEM` is then simulated with the calibrated rates to generate estimated daily cases and compared against reported confirmed cases. The confirmed daily positive (*y*^+^) and daily negative (*y*^−^) were obtained from public sources (CTW nd) between March 22, 2020 to March 1, 2021. Forecasts were made for a 7-day period after each weekly calibration of the rates.

Figures 2a and 2b show that `PySIRTEM` is quite accurate in capturing the trends of infection for Arizona. Figure 2c displays the intrinsic parametric trends which explain the daily pandemic progression i.e., the peak in *β* at week 10, 30, and 40 correspond with the following rise in positive cases. In Figure 2d, we compare the mean daily error in estimation for 20 iterations from `SIRTEM`’s AR(2) calibration method (Azad et al. 2022) and `PySIRTEM`’s particle filter approach, with `PySIRTEM` providing more accurate results. Comprehensive error comparisons for other states are listed in Table 2.

**Table 2.**
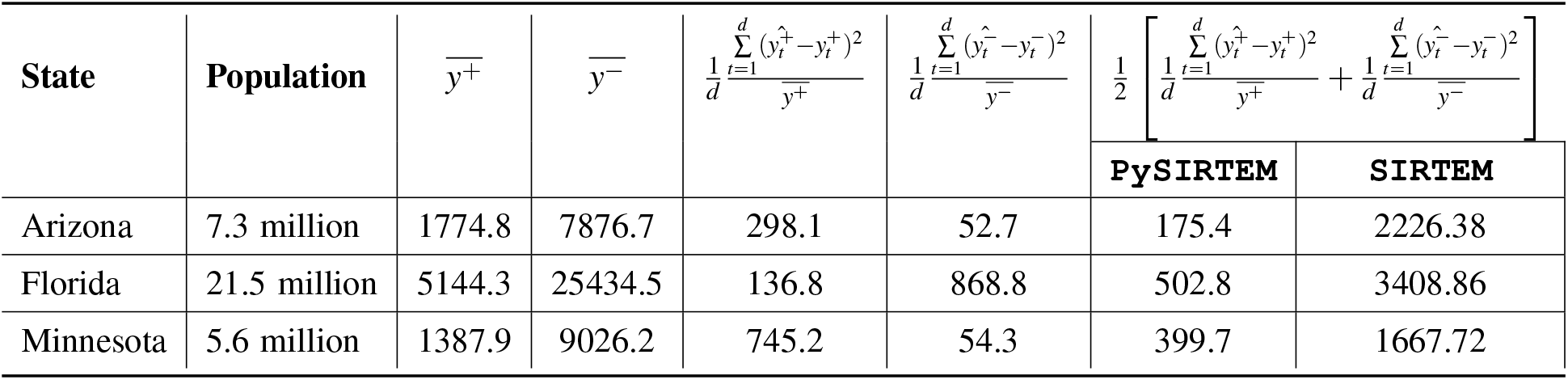
Normalized Mean-squared Error for selected U.S States from PySIRTEM vs SIRTEM Calibration results.

**Figure 2.**
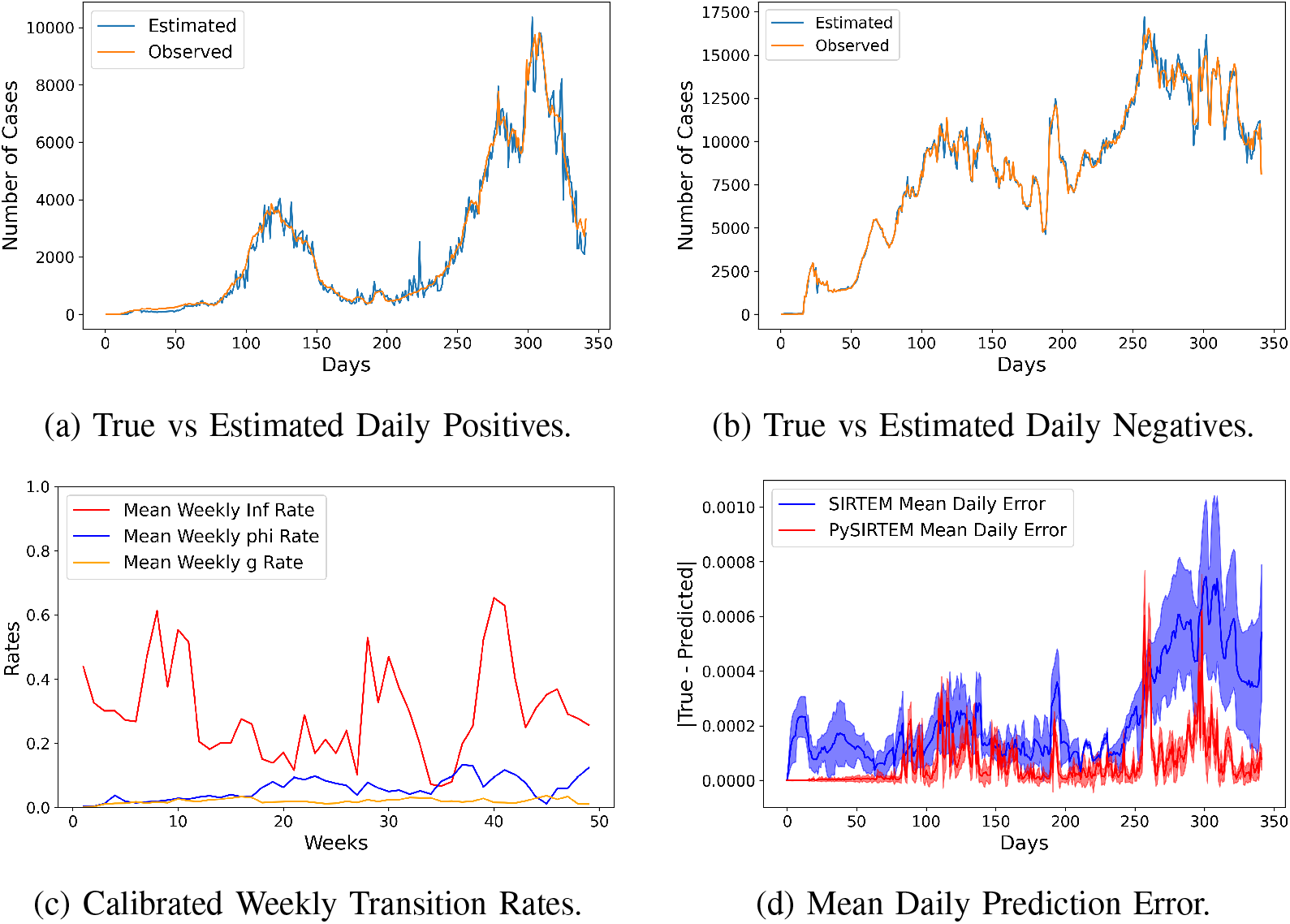
Model prediction results for the State of Arizona.

### 3.2. Factorial Analysis Outcomes

In this section, we analyze how different infection rate (*β*), testing rate (*φ*), and general sickness rate (*g*) impact daily case numbers; in particular, we aggregate the findings from factorial analysis in pairplots in Figure 3. For each of the experiments, we keep the infection rate trends constant, only changing the testing rate and general sickness rate trends to comparing the pairwise interaction between the factors. The diagonal plots in the matrices denote the main effects while the off-diagonal plots indicate interaction. At each cellplot at row *r* and column *c*, the red line refer to relation between factor *c* and the peak value (maximum daily positive cases reported) when factor *r* is not used in the simulation and the blue line refers to the interaction between factor *c* and the peak value in the presence of factor *r*.

**Figure 3.**
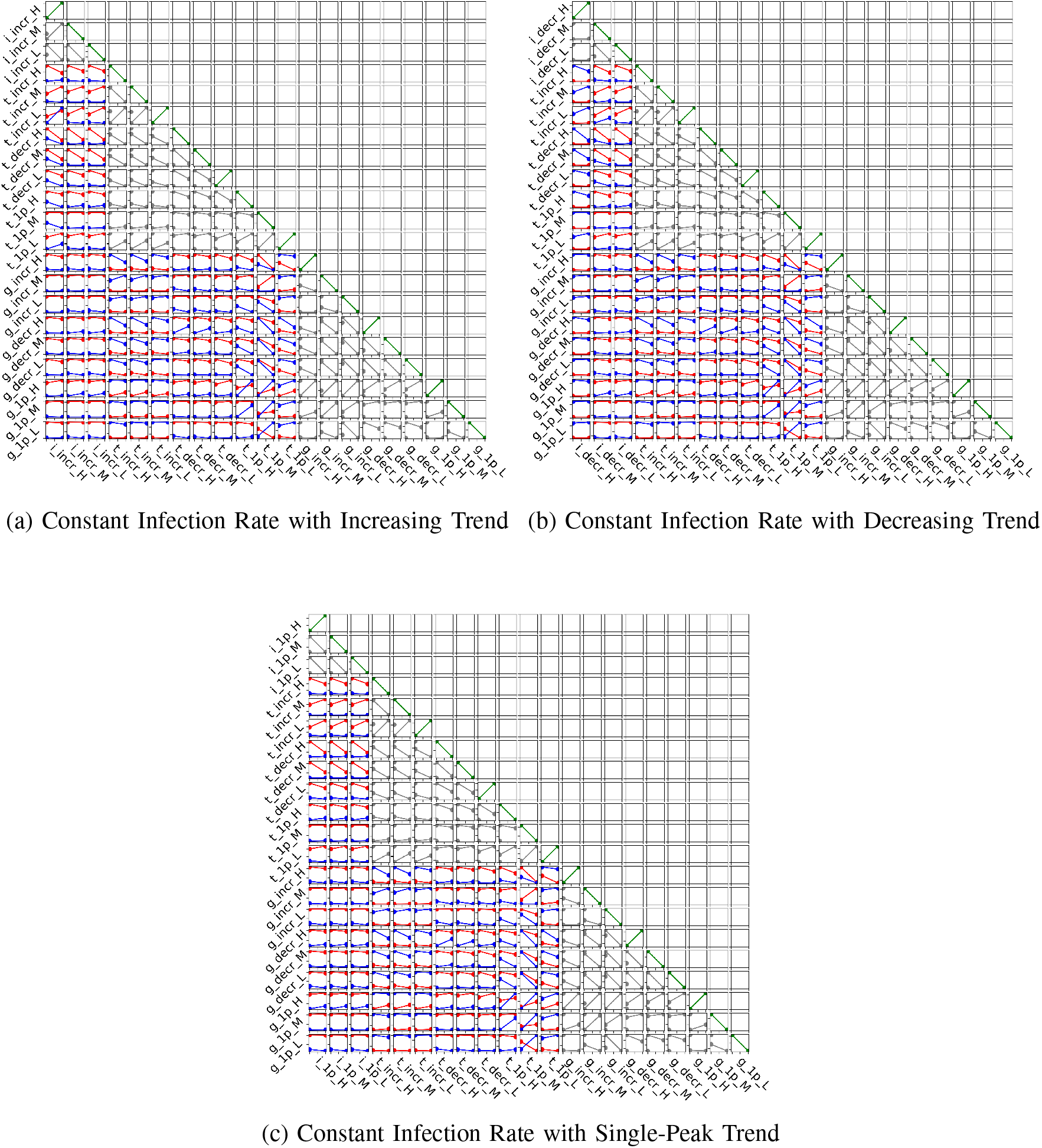
Factorial Analysis for different trends in `PySIRTEM` transition parameters (Outcome variable: maximum daily positive cases). Factors in *x* and *y*-axis are abbreviated as follows: *i, t, g* for infection, testing, and general sickness rate respectively; *incr, decr*, and 1*p* for increasing, decreasing, and single-peak trends; *H, M*, and *L* for the rate bounds (high, medium, low).

For space scarcity, factor names are abbreviated in Figure 3. For example, *i*_*incr*_*H* refers to high increasing infection rate trend. The values for each trend are again generated from Table 1. Since no two trends for the same factor are possible for an independent run of the simulation, interaction plots for same factors have been grayed out.

In the plots in Figure 3a, we see non-parallel lines between increasing *β* and both increasing and decreasing *φ*, implying strong levels of interaction among these. Specifically, when *φ* increases, more positive cases are detected, suggesting higher testing is effective in identifying infections (*t*_*incr*_*M, i*_*incr*_*M*). However, decreasing *φ* leads to under-reporting (*t*_*decr*_*H, i*_*incr*_*H*) as the lines point downward (lower positive cases) even when *β* is increasing. We can observe that a testing policy where majority of testing is concentrated on a single timepoint (*t*_*incr*_*M*) is less effective in containing spread, as implied by the parallel lines (*t*_1*p*_*M, i*_*incr*_*M*).

The plots in Figure 3a further show that with increasing *β*, non-COVID *g* rate has trivial impact on the peak of infection because the number of false positives due to *g* rate is still greatly outnumbered by true positives (*g*_*incr*_*M, i*_*incr*_*M*). The *g* rate, however, shows strong interaction with a single-peak testing rate policy. Particularly, when a high number of individuals are sick with non-COVID illness, high testing rate may lead to more false positives stimulating higher positive reporting (*g*_1*p*_*H, t*_1*p*_*H*). This is evident from the blue line having a steeper positive slope than the red even when *β* trend is constant in both cases.

Figure 3b displays that if *β* is decreasing rapidly, a high test rate can quickly flatten the infection curve resulting in lower positive cases (*t*_*incr*_*H, i*_*decr*_*H*). However, a low increasing *φ* may not be sufficient to curb a slowly decreasing infection trend (*t*_*incr*_*L, i*_*decr*_*H*), stressing the significance of adaptive testing policies. Here the blue line displays positive slope implying insufficient testing which leads to higher spread and higher reporting of positive cases.

Figure 3c reveals that for bell-curve *β* trend, a similar single-peak testing policy is mostly ineffective as indicated by parallel interaction plots for (*t*_1*p*_*M, i*_1*p*_*M*). However, increasing and decreasing test rates are capable of containing positive cases. Moreover, plot for (*t*_*decr*_*M, i*_1*p*_*M*) shows that decreasing *φ* (i.e. strong testing at the start) is more effective in keeping the reported cases low (the downward slope suggests lower spread and reporting) than increasing *φ* (*t*_*incr*_*M, i*_1*p*_*M*) where we see a positive slope which demonstrates that testing policy failed to regulate infection.

### 3.3. Containment Scenario Analysis

In order to demonstrate the capabilities of `PySIRTEM` as a simulation tool, we conduct a series of experiments comparing results generated from parameter setups displaying low and strong interaction derived from factor analysis. We gauge the rate of impact when different rates change their trends. The outcomes are compared against the simulation results obtained from the setup where the rates display only one trend i.e. increasing or decreasing. Finally, we evaluate the rate of change in infection and positive cases in order for better policy-making.

We run the simulation for *N* = 150 days and the trends are kept constant after day 100 to observe the effect of the trends. At *n* = 75, we toggle the trends of one of the rates. For example, testing rate (*φ*) follows a decreasing trend up to day 75 and then it gradually starts increasing in Figure 4a. In each of the experiments, we keep the other rates’ trends unchanged i.e., the infection (*β*) and general sickness rate (*g*) follow an increasing trend throughout. Similarly, when *β* changes trends, *φ* and *g* are kept as increasing. The positive (*y*^+∗^) and infected (*I*^∗^) case numbers are compared against data obtained from `PySIRTEM` where the rates have unchanged trends i.e *β* always increasing and *φ* always decreasing. The rates of change are calculated as follows,

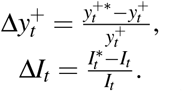

As we see in Figure 4a, when *φ* starts to increase (after the vertical red dashed line), there is a quick surge in reported cases; however the infected numbers gradually decrease and the higher testing not only limits the peak of infection but also delays it. On the other hand, Figure 4b shows that, when *β* suddenly starts to increase, the infected hill is wider even though the testing is able to identify the peak; this suggests that infection is lasting and stronger contingency plans are required. These experiments, along with others involving various hyperparameters, give valuable insights in managing resources and policy making during different scenarios during the course of a pandemic.

**Figure 4.**
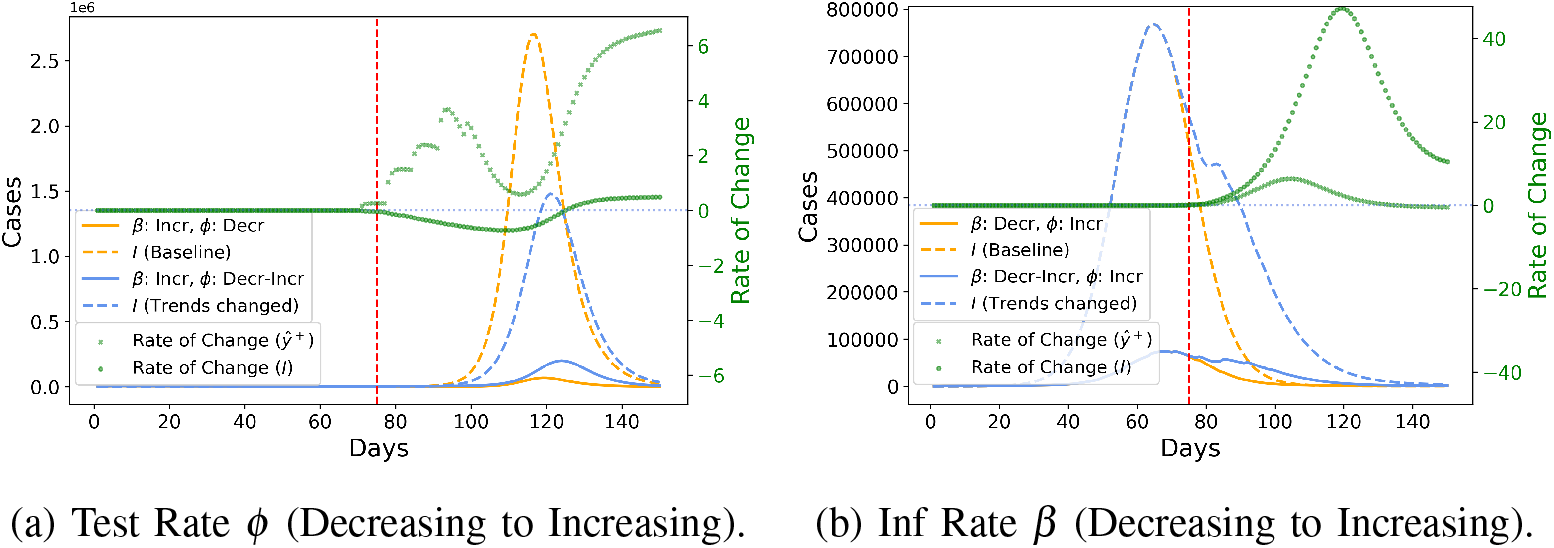
Comparison of Model prediction results for two different scenarios.

## 4. CONCLUSION

`PySIRTEM` provides a powerful and flexible tool for simulating pandemic dynamics via the addition of a particle filter for dynamic estimation of parameters. Our factorial design analysis showcases that `PySIRTEM` is able to capture critical epidemiological factor interactions, including infection and test rates, and facilitates informed decision-making. The capability of the model to simulate various real world pandemic scenarios demonstrates its versatility. Future efforts could see us explore additional variables like vaccination techniques, mutated viral strains, and demographic traits in the context of `PySIRTEM`. To further enhance granularity, an agent-based model (ABM) can be added to simulate behavior and interaction at the individual level to supplement `PySIRTEM`’s population-level dynamics.

## Data Availability

All data produced in the present study are available upon reasonable request to the authors

## AUTHOR BIOGRAPHIES

**PREETOM BISWAS** is an Undergraduate Student at Arizona State University for the School of Computing and Augmented Intelligence (SCAI) working with Dr. Giulia Pedrielli and Dr. K. Selçuk Candan on Pandemics modeling. His research focuses on stochastic simulation and optimization within with epidemiology applications. His email address is.

**GIULIA PEDRIELLI** is currently Associate professor for the School of Computing and Augmented Intelligence (SCAI) at Arizona State University. She develops her research in design and analysis of random algorithms for global optimization, with focus on improving finite time performance and scalability of these approaches. Applications of her work are in individualized cancer care, bio-manufacturing, design and control of self-assembled RNA structures, verification of cyber-physical systems. Her email address is and her website is https://www.gpedriel.com/.

**K. SELÇUK CANDAN** is a professor of computer science and engineering at Arizona State University (ASU) and the director of ASU’s Center for Assured and Scalable Data Engineering (CASCADE). His primary research interest is in the area of management and analysis of non-traditional, heterogeneous, and imprecise (such as multimedia, web, and scientific) data, with applications in public heath and sustainability, among others. He is an ACM Distinguished Scientist. His email address is and his website is https://kscandan.site.

## Notes

### Competing Interest Statement

The authors have declared no competing interest.

### Funding Statement

This study was funded by NSF#2335670

